# Testing a polygenic risk score for morphological microglial activation in Alzheimer’s disease and aging

**DOI:** 10.1101/2023.03.10.23287119

**Authors:** Earvin S. Tio, Timothy J. Hohman, Milos Milic, David A. Bennett, Daniel Felsky, the Alzheimer’s Disease Neuroimaging Initiative

## Abstract

Neuroinflammation and the activation of microglial cells are among the earliest events in Alzheimer’s disease (AD). However, direct observation of microglia in living people is not currently possible. Here, we indexed the heritable propensity for neuroinflammation with polygenic risk scores (PRS), using results from a recent genome-wide analysis of a validated post-mortem measure of morphological microglial activation. We sought to determine whether a PRS for microglial activation (PRS_mic_) could augment the predictive performance of existing AD PRSs for late-life cognitive impairment. First, PRS_mic_ were calculated and optimized in a calibration cohort (Alzheimer’s Disease Neuroimaging Initiative (ADNI), n=450), with resampling. Second, predictive performance of optimal PRS_mic_ was assessed in two independent, population-based cohorts (total n=212,237). Our PRS_mic_ showed no significant improvement in predictive power for either AD diagnosis or cognitive performance. Finally, we explored associations of PRS_mic_ with a comprehensive set of imaging and fluid AD biomarkers in ADNI. This revealed some nominal associations, but with inconsistent effect directions. While genetic scores capable of indexing risk for neuroinflammatory processes in aging are highly desirable, more well-powered genome-wide studies of microglial activation are required. Further, biobank-scale studies would benefit from phenotyping of proximal neuroinflammatory processes to improve the PRS development phase.

## Introduction

Neuroinflammation is an important process in the pathogenesis of Alzheimer’s disease (AD)^1–5^. Efforts to identify inflammatory biomarkers for early diagnosis or improved prognosis of AD have largely been focused on measuring signaling proteins in blood and cerebrospinal fluid (CSF).^6,7^ In the central nervous system, the activation of microglia - the brain’s resident immune cells - is an important mediator of inflammation in AD. The process of activating microglia from their “resting”, or sentinel, state is closely tied to immune signaling, and enables the removal of potentially toxic protein aggregates (such as Aβ) via phagocytosis.^8^ Dysregulated immune signaling, such as in the case of chronic neuroinflammation, may lead to further microglial activation, which has been shown to form a feedback loop with amyloid deposition, with both processes exacerbating the other.^3^

Determining the activation states of microglia in humans is an active field. Recent work from our group has established a post-mortem morphological phenotype, termed the proportion of activated microglia (PAM), which is strongly associated with AD neuropathology, rates of cognitive decline in aging, and neuroimmune gene expression signatures from isolated human microglia.^9,10^ While this measure can proxy for microglia-mediated neuropathological cascades in post-mortem studies of aging and AD, the inaccessibility of human brain tissue in living people poses important limitations on the utility of PAM. Given its important etiological role in AD, measuring the propensity for microglial activation in living people over the course of aging may be a useful way of stratifying at-risk populations. Further, such a stratification would provide mechanistic information, and therefore could be useful for implementing precision treatment strategies. Unfortunately, existing methods for determining microglial activation states *in vivo* require expensive and invasive imaging protocols using positron emission tomography (PET).^11^

To develop a minimally invasive and inexpensive tool for identifying individuals at risk for AD based on their propensity for morphological microglial activation *in vivo*, we turned to genetic markers measurable in saliva or blood. Using genome-wide methods, we previously identified risk loci for the PAM phenotype in the inferior temporal (IT) and midfrontal (MF) cortices.^10^ These genetic maps can be used for the calculation of polygenic risk scores (PRS), which are commonly used to bridge the gap between genetic research and clinical utility by summarizing an individual’s genetic susceptibility for disease based on genotype information.^12,13^ PRSs have been deployed in several diseases, notably in cardiovascular diseases,^14–16^ including Alzheimer’s disease.^17–19^ Despite the large effect sizes of the single nucleotide polymorphisms (SNPs) in the *APOE* region, inclusion of more SNPs in AD PRSs has shown to improve predictive power.^20–22^ However, most PRSs developed to date use as their source case-control studies on AD diagnosis, while biologically-informed PRSs are lacking.

Our primary goal for this study was to develop and validate a novel PRS for microglial activation (PRS_mic_) and assess the capacity of this PRS_mic_ to augment the predictive performance of existing PRS for AD (PRS_AD_). We hypothesized that models including this PRS_mic_ will improve predictive performance over models with PRS_AD_ alone, since existing gold-standard PRS_AD_ are based on clinically defined Alzheimer’s dementia and may be missing crucial etiological information related to neuroinflammatory mechanisms. Our secondary goal was to explore any putative associations PRS_mic_ with specific AD-related neuropathologies *in vivo*, including beta-amyloid, hyperphosphorylated tau, and inflammatory AD-related biomarkers (tumor necrosis factor alpha, TNF-α, and neurofilament light chain, NfL) measured in both brain (PET) and periphery (blood and CSF). In order to adhere to the highest standards of rigor and transparency in PRS development and reporting, we followed the methodological framework described by the Clinical Genome Resource (ClinGen) Complex Disease Working Group and the Polygenic Score (PGS) Catalog Polygenic Risk Score Reporting Standards (PRS-RS).^23^

## Materials and methods

### Study datasets and outcomes

#### The Alzheimer’s Disease Neuroimaging Initiative (ADNI)

Data used in the preparation of this article were obtained from the Alzheimer’s Disease Neuroimaging Initiative (ADNI) database (adni.loni.usc.edu). The ADNI was launched in 2003 as a public-private partnership, led by Principal Investigator Michael W. Weiner, MD. For up-to-date information, see www.adni-info.org. The ADNI is a longitudinal study consisting of four study phases (ADNI-1, ADNI-GO, ADNI-2, and ADNI-3). Participants are enrolled in a case-control design (cognitively normal (CN), significant memory concern (SMC), early mild cognitive impairment (EMCI), late mild cognitive impairment (LMCI) and clinical Alzheimer’s disease (AD)) and clinical, imaging, fluid biomarker, and genetic data are collected at baseline and follow-up (6- and 12-month).^24^ In total, we analyzed up to n=973 participants (50.8% with an AD diagnosis) for PRS calculation and hyperparameter tuning, and n=1,404 participants for exploratory analyses with multiple AD biomarkers and neuropathologies. Informed written consent was obtained by the ADNI investigators at each participating ADNI site. Descriptive statistics of the variables included in this study can be found in **Table S1**.

Our primary outcome used for calibration of PRS_mic_ was tumour necrosis factor alpha (TNF-α) measured in plasma using the Luminex xMAP platform by Rules-Based Medicine (RBM). Plasma levels of TNF-α have been previously shown to contribute partially to an inflammatory factor that predicts decline in executive function in patients with mild Alzheimer’s disease.^25^ A total of n=450 participants had at least one measure of TNF-α available. For participants with repeat measures, an average of the measurements was used as the outcome. For the calibration of PRS_AD_, we derived a subset of only participants in the cognitively normal (n=479) and Alzheimer’s disease (n=494) categories. For exploratory analyses, we did not restrict participants by diagnostic category. Primary outcomes for these analyses include the Alzheimer’s Disease Assessment Scale–Cognitive Subscale (ADAS-Cog), where a higher score indicates greater cognitive impairment. Pathological outcomes were also analyzed, including peripheral measures of known AD-related and inflammatory biomarkers such as amyloid, tau, phosphorylated tau, and neurofilament light chain (NfL) in CSF. Levels of NfL in plasma were also analyzed. Brain-based measures of amyloid and tau were derived from standardized uptake value ratios (SUVR) of PET tracers ^18^F-AV-45 and ^18^F-AV-1451, standardized to the whole cerebellum and the inferior cerebellar reference region, respectively. These measures were localized to the inferior temporal and midfrontal regions of the brain, averaged across both hemispheres.

Genotype data for participants were obtained at every phase: n_ADNI-1_=757 genotyped using the Illumina Human610-Quad BeadChip, n_ADNI-GO/2_=432 genotyped using the Illumina HumanOmniExpress BeadChip, and n_ADNI-3_=327 genotyped using the Illumina Global Screening Array v2. Standard quality control was applied to each set of genotypes separately before imputation was performed on the TOPMed Imputation Server, resulting in 8,028,924 high-quality variants in n=1,569 participants.

#### The UK Biobank

The UK Biobank is a large-scale biomedical database of over half a million participants residing throughout the United Kingdom.^26^ Volunteers were initially enrolled over a four year period beginning in 2006, aged 40 to 69, and will be followed with either repeat visits or questionnaire data to track their health outcomes. Linked electronic health records through the National Health Service also provided insight into the participants’ health, notably giving access to International Classification of Disease (ICD-10) codes from in-patient health records. We analyzed up to n=200,924 elderly participants of the UK Biobank aged 60 and over at time of recruitment. All participants provided written informed consent to the UK Biobank. Descriptive statistics of the variables included in this study can be found in **Table S1**.

The primary study outcome measure utilized in the UK Biobank is the presence of an International Classification of Disease (ICD-10) G30 code for Alzheimer’s disease. These codes were ascertained from any of the following: death register, primary care records, or hospital admission data, and binarized into presence or absence of at least one record for each individual in our study. Genotype data were derived from blood samples collected at the initial assessment: n=487,442 genotyped using the Applied Biosystems UK Biobank Axiom Array. After quality control, 670,739 autosomal markers remained.^26^ Finally, 93,095,623 autosomal variants were imputed using the Haplotype Reference Consortium^27^ and UK10K + 1000 Genomes reference panels.^28^ We used the UK Biobank as one of our external test cohorts.

#### The Canadian Longitudinal Study on Aging (CLSA)

CLSA is a longitudinal, nation-wide study of Canadians aimed at investigating the aging process from mid-through late-life (aged 45 to 85 at recruitment), with 20 year follow-up.^29^ Data were collected on approximately 50,000 randomly selected Canadians via computer-assisted telephone interviews, including measures of cognition, sociodemographics, health equity, amongst others. Additionally, a subset of approximately 30,000 participants underwent further examination at select data collection sites and provided biospecimen samples for genotyping. We analyzed up to n=11,313 elderly participants of the CLSA aged 60 and over at time of recruitment. All participants provided informed written consent to the CLSA. Descriptive statistics of the variables included in this study can be found in supplementary **Table S1**.

Our primary evaluated outcome measure in CLSA was a latent score of overall cognition, adjusted for age, sex, and education status. This composite score is constructed from participant performance on the following cognitive tests: the Rey Memory Test, the Animal Fluency Test, the Mental Alternation Test, the Verbal Fluency Test (FAS Test), and the Stroop Test, standardized to a mean of 100 and a standard deviation of 15.^30^ A higher overall cognition score indicates better cognitive performance. Genotyping was performed using the Axiom 2.0 Assay Automated Workflow on Affymetrix NIMBUS protocol.^31^ Samples were also hybridized to UK Biobank arrays and 794,409 genetic variants survived quality control. Imputation was conducted with TOPmed reference panels, resulting in genotypes for approximately 308 million genetic variants.^31^ We used CLSA as our second external test cohort.

### GWAS summary statistics for PRS calculation

To develop our PRS_mic_, we accessed summary statistics from our published genome-wide association study (GWAS) of the proportion of active microglia (PAM) phenotype measured in post-mortem brain samples from the Religious Orders Study and Rush Memory and Aging Project (ROS/MAP) cohorts.^10,32^ The PAM phenotype provides an index of microglial activation, measured by HLA staining and manual counting of morphologically activated microglia, relative to basal levels of total microglia (including “inactive”, or sentinel microglia) within a brain region.^10,33^ Specifically, we retrieved genome-wide summary statistics from two GWAS of PAM quantified in two different brain regions: 1) inferior temporal (IT) cortex and 2) midfrontal (MF) cortex. These regions were chosen as they were the only regions linked to Alzheimer’s disease neuropathology and cognition in previous work.^10^ For the calculation of our baseline PRS_AD_, we used summary statistics from the largest and most recent GWAS on late-onset AD and related dementias.^34^

### PRS calculation

The clumping & thresholding (C+T) method^35^ implemented in PRSice-2^36^ was used to calculate all PRSs in our study. This method requires the selection of three hyperparameters: 1) linkage disequilibrium (LD) clumping squared correlation (r^2^) threshold, 2) LD clumping window size, and 3) *p*-value threshold for SNP inclusion. The PRS is then calculated as the effect-weighted (with weights equal to beta coefficient from the source GWAS) sum of allelic dosage across included SNPs, per individual. Given the strong influence of hyperparameters on the performance of downstream C+T PRSs, we performed parameter optimization across a large set of values according to published guidelines^37^:

- LD clumping r^2^ = {0.01, 0.05, 0.1, 0.2, 0.5, 0.8, 0.95}.
- LD clumping window base size = {25, 50, 100, 200, 500} (In PRSice, the window size is then computed as the base size divided by the correlation threshold).
- A sequence of 605 SNP inclusion thresholds (100 per order of magnitude, ranging from genome-wide significance of 5×10^−8^ to 0.05), as well as {0.1, 0.2, 0.5, 0.8, 1}.

Across all combinations of parameters, PRSs composed of less than 10 SNPs and duplicate scores were discarded. The most conservative parameters were used to select which of the duplicate scores to retain—taking the smallest SNP-inclusion and LD clumping r^2^ thresholds, and the largest clumping window size in that order of priority. Finally, to account for fine population structure, the first ten genetic principal components were regressed from each score, yielding standardized residuals that were carried forward for analysis.

### Statistical analysis

Analyses proceeded in three phases: 1. ***Calibration***: we calculated a series of PRS_mic_ in our calibration sample (ADNI) and performed optimal clumping and thresholding hyperparameter selection using bootstrap resampling. 2. ***Validation***: we assessed the performance of models including our optimized PRS_mic_ in elderly participants from two independent, population-based cohorts. We also tested differences in performance of models including only established AD PRS vs. models including our novel PRS_mic_ + PRS_AD_. 3. ***Exploration***: following steps 1 and 2, designed to meet our first study goal, we relaxed statistical correction thresholds to identify putative associations of PRS_mic_ with a set of *in vivo* AD biomarkers within the deeply-phenotyped ADNI sample.

#### Calibration of microglial PRS (PRS_mic_) in ADNI

We first calculated two sets of PRS_mic_ in the non-Hispanic white ADNI subsample (n=1,414), corresponding to the two source GWAS performed for PAM in two brain regions; we refer to these scores as PRS_mic_[MF] for midfrontal cortex, and PRS_mic_[IT] for inferior temporal cortex. Logistic regression models were fit to test the associations of the PRS_mic_ with levels of plasma tumor necrosis factor alpha (TNF-α) in *n*=450 samples with complete data. In the absence of a comparable morphological microglial activation phenotype in any existing datasets outside of the ROS/MAP samples that were used in the source PAM GWAS, TNF-α was used as a proxy for microglial activation in ADNI due to the known phenomenon of TNF-α-mediated microglia overactivation.^38^ We searched a parameter space of seven clumping correlation thresholds, five clumping window sizes, and 605 SNP-inclusion thresholds for a total of 21,175 parameter combinations. After ensuring >10 SNPs were included in a score and duplicate scores were removed, we were left with a total of 9,685 PRS_mic_[IT] and 9,228 PRS_mic_[MF] scores for optimization. We fit separate linear models for each score, covarying for age, sex, and years of education, then performed 1,000 iterations of bootstrap resampling on every model. Bootstrapped *r*^*2*^ were obtained for each model and the top performing combinations of hyperparameters (according to median bootstrapped *r*^*2*^) were selected and used to calculate PRS_mic_ for modelling in CLSA and UK Biobank.

To calibrate our PRS_AD_ for downstream modelling and comparison, we performed a similar procedure but using clinical AD diagnosis at last study visit as the outcome (*n*=973, 49.2% cognitively normal control, excluding MCI cases), including the same covariates. For each bootstrap sample, a model was trained on 70% of the data (*n*=681) and AUC calculated using the remaining 30% test set (*n*=292). Optimal construction parameters were selected based on median bootstrapped AUC. The bootstrap selection procedure was then repeated without the inclusion of covariates in the bootstrapped models, as a point of comparison.

#### Testing the predictive performance of PRS_mic_ for AD diagnosis in UK Biobank and cognitive performance in CLSA

PRS_mic_ (IT and MF) and PRS_AD_ were calculated using calibrated hyperparameters in both UK Biobank and CLSA and tested for predictive performance in models of AD diagnosis and cognitive function, respectively. To determine the improvement in models with the addition of PRS_mic_, we compared 1) baseline clinical covariate-only models, 2) baseline models plus an additional PRS_AD_ term, and 3) fully augmented models with both PRS_AD_ and PRS_mic_ terms added. We calculated changes in AUC (for binary AD diagnosis) and changes in *r*^*2*^ (for continuous cognitive scores) resulting from the inclusion of the PRS_mic_ term. Likelihood ratio tests (LRT) were performed to assess if fully augmented models represent an improvement over baseline models. Biological sex, age, and level of education (using the International Standard Classification for Education (ISCED) definitions in the UK Biobank)^39^ were also included as covariates in the predictive models.

#### Exploration of PRS_mic_ associations with in vivo AD biomarkers in ADNI

Following the calibration and validation of PRS_mic_ in UK Biobank and CLSA, we returned to the ADNI calibration sample to map exploratory associations between PRS_mic_ and several pathological and cognitive variables *in vivo*. Multiple testing correction was performed using the Bonferroni method across the number of tested phenotypes and both PRS_mic_ derivations (i.e. IT and MF; critical *p*=0.05/11/2=0.0023), but no correction was performed within PRS across construction parameters. Given the exploratory aim of this part of the analyses, we deemed this more liberal correction approach suitable. Models included biological sex, age, and education level as covariates.

## Results

### Calibration of PRS_mic_ against plasma TNF-α in ADNI

Results from the bootstrap model selection procedure showed variability in model performance according to hyperparameter combinations regardless of covariate inclusion (**Figure S1**) or exclusion (**Figure S2**) in the calibration process. The optimal combination of hyperparameters for each PRS are shown in **Table 1**. In ADNI, these optimized PRS_mic_ were significantly associated with levels of TNF-α (PRS_mic_[IT]: β=0.101, *p*=0.026; PRS_mic_[MF]: β=-0.126, *p*=5.25×10^−3^) and the PRS_AD_ was strongly predictive of AD diagnosis (β=0.592, *p*=4.68×10^−15^). Importantly, these models are not unbiased indicators of performance, as they are overfit due to sample overlap. Also, we observed that the effect for the midfrontal score was inconsistent with the other scores, suggesting that genetically-determined greater propensity for morphological microglial activation in midfrontal cortex might be associated with lower levels of circulating TNF-α.

**Table 1:** Optimal PRS construction parameters selected from the bootstrap selection procedure.

|  | Calibration Model | Squared correlation threshold | Window size | SNP inclusion threshold | # of SNPs in PRS | Maximal bootstrapped metric* | Bootstrap 95% CI |
| --- | --- | --- | --- | --- | --- | --- | --- |
| PRS<sub>mic</sub>[IT] | With covariates | 0.1 | 5,000 | 0.1 | 47,158 | 0.048 | [0.02,0.09] |
|  | Without covariates | 0.1 | 5,000 | 0.1 | 47,158 | 0.010 | [3.05x10<sup>-4</sup>, 0.036] |
| PRS<sub>mic</sub>[MF] | With covariates | 0.2 | 125 | 2.75x10<sup>-5</sup> | 27 | 0.054 | [0.02,0.10] |
|  | Without covariates | 0.2 | 125 | 3.02x10<sup>-5</sup> | 29 | 0.015 | [8.26x10<sup>-4</sup>, 0.045] |
| PRS<sub>AD</sub> | With covariates | 0.1 | 250 | 3.65x10<sup>-6</sup> | 251 | 0.675 | [0.62,0.73] |
|  | Without covariates | 0.2 | 125 | 1.49x10<sup>-6</sup> | 307 | 0.621 | [0.56,0.68] |

\*Area under the ROC curve (AUC) was used for Alzheimer's disease diagnosis (with a 70/30 training/testing split) and variance explained ( $r^2$ ) for TNF-α (trained on the entire sample). The upper row for each PRS are statistics derived from regression models including covariates (age, sex, and years of education), and the lower for scores excluding covariates.

### Testing of PRS_mic_ in UK Biobank and CLSA

First, we tested the predictive performance of each optimized PRS individually on AD diagnosis (UK Biobank) and cognitive performance (CLSA), with and without the addition of covariates (results are summarized in **Table 2**). For modelling of AD diagnosis in UK Biobank, the addition of PRS_AD_ significantly improved baseline covariates-only models (*p*=1.2×10^−237^; ΔAUC=0.07), as expected (**Table 3**). Testing the addition of PRS_mic_ into baseline covariates-only models, however, did not result in any significant improvements (IT: *p*=0.77; MF: *p*=0.45). Similarly, for modelling of cognitive performance in CLSA, the addition of PRS_AD_ improved baseline covariates-only models (Δ*r*^*2*^=5.97×10^−4^, *p*=0.01), while PRS_mic_ did not (IT: *p*=0.96; MF: *p*=0.82) (**Table 3**). These results suggest that, on their own, the PRS_mic_ do not hold significant predictive power for AD diagnosis or cognitive performance in the general mid-late life population.

**Table 2:** Estimated effect and significance of PRS terms in linear models of Alzheimer’s disease diagnosis and cognitive functioning.

|  | Calibration Model | PRS<sub>mic</sub>[IT] | PRS<sub>mic</sub>[MF] | PRS<sub>AD</sub> |
| --- | --- | --- | --- | --- |
| UK Biobank (AD diagnosis, AUC) | With covariates | -5.33x10<sup>-3</sup> (0.78) | 0.011 (0.57) | **0.603 (5.66x10<sup>-249</sup>)** |
|  | Without covariates | -8.69x10<sup>-3</sup> (0.65) | 8.73x10<sup>-3</sup> (0.65) | **0.572 (2.63x10<sup>-207</sup>)** |
| CLSA (cognitive score, $r^2$) | With covariates | 2.48x10<sup>-3</sup> (0.79) | 1.68x10<sup>-3</sup> (0.86) | **-0.021 (0.03)** |
|  | Without covariates | 5.55x10<sup>-3</sup> (0.55) | 1.64x10<sup>-3</sup> (0.86) | -0.014 (0.15) |

The upper rows for each cohort are statistics derived from scores calibrated in ADNI with covariates (age, sex, and years of education), and the lower for scores calibrated without covariates. $p$ -values are calculated from regression models, with significant terms in bold.

**Table 3:** Change in model performance with inclusion of different polygenic risk scores.

| | | ΔAUC/Δ$r^2$ compared to “covariates-only” models ($p$-value) | | | ΔAUC/Δ$r^2$ compared to “covariates + PRS<sub>AD</sub>” models ($p$-value) | |
| --- | --- | --- | --- | --- | --- | --- |
|  |  | PRS<sub>mic</sub>[IT] | PRS<sub>mic</sub>[MF] | PRS<sub>AD</sub> | PRS<sub>mic</sub>[IT] | PRS<sub>mic</sub>[MF] |
| UK Biobank (AD diagnosis, ΔAUC) | With covariates | -9.57x10<sup>-5</sup> (0.77) | 1.02x10<sup>-4</sup> (0.45) | **6.66x10<sup>-2</sup> (1.24x10<sup>-237</sup>)** | -4.83x10<sup>-5</sup> (0.58) | 2.04x10<sup>-4</sup> (0.39) |
|  | Without covariates | -1.53x10<sup>-4</sup> (0.60) | -8.14x10<sup>-5</sup> (0.52) | **5.59x10<sup>-2</sup> (9.55x10<sup>-202</sup>)** | 1.26x10<sup>-4</sup> (0.40) | 4.71x10<sup>-5</sup> (0.46) |
| CLSA (cognitive score, Δ$r^2$) | With covariates | 2.30x10<sup>-7</sup> (0.96) | 4.37x10<sup>-6</sup> (0.82) | **5.97x10<sup>-4</sup> (0.01)** | 5.14x10<sup>-7</sup> (0.94) | 5.33x10<sup>-6</sup> (0.80) |
|  | Without covariates | 1.53x10<sup>-5</sup> (0.67) | 4.84x10<sup>-6</sup> (0.81) | 3.08x10<sup>-4</sup> (0.06) | 1.60x10<sup>-5</sup> (0.67) | 5.92x10<sup>-6</sup> (0.79) |

The upper rows for each cohort are statistics derived from scores calibrated in ADNI with covariates (age, sex, and years of education), and the lower for scores calibrated without covariates. $p$ -values are calculated from likelihood-ratio tests, with significant results (in bold) indicating a better statistical fit with the inclusion of the extra term tested.

We then tested the ability of PRS_mic_ to improve models of AD diagnosis and cognitive performance over and above models including PRS_AD_. Augmenting models of cognition that also include PRS_AD_ terms with PRS_mic_ did not significantly improve them in either UK Biobank or CLSA (**Figure 1**). We also tested whether hyperparameters optimized using covariate-inclusive or -exclusive models altered performance in any important ways. Results were largely unchanged when analyses were repeated with scores calibrated controlling for age, sex, and education levels (**Figure S3**). One minor observation was that PRS_AD_ scores calibrated without covariates included showed less significant improvement of covariate-only models of cognitive performance in CLSA (Δ*r*^*2*^=3.08×10^−4^, *p*=0.06).

**Figure 1:**
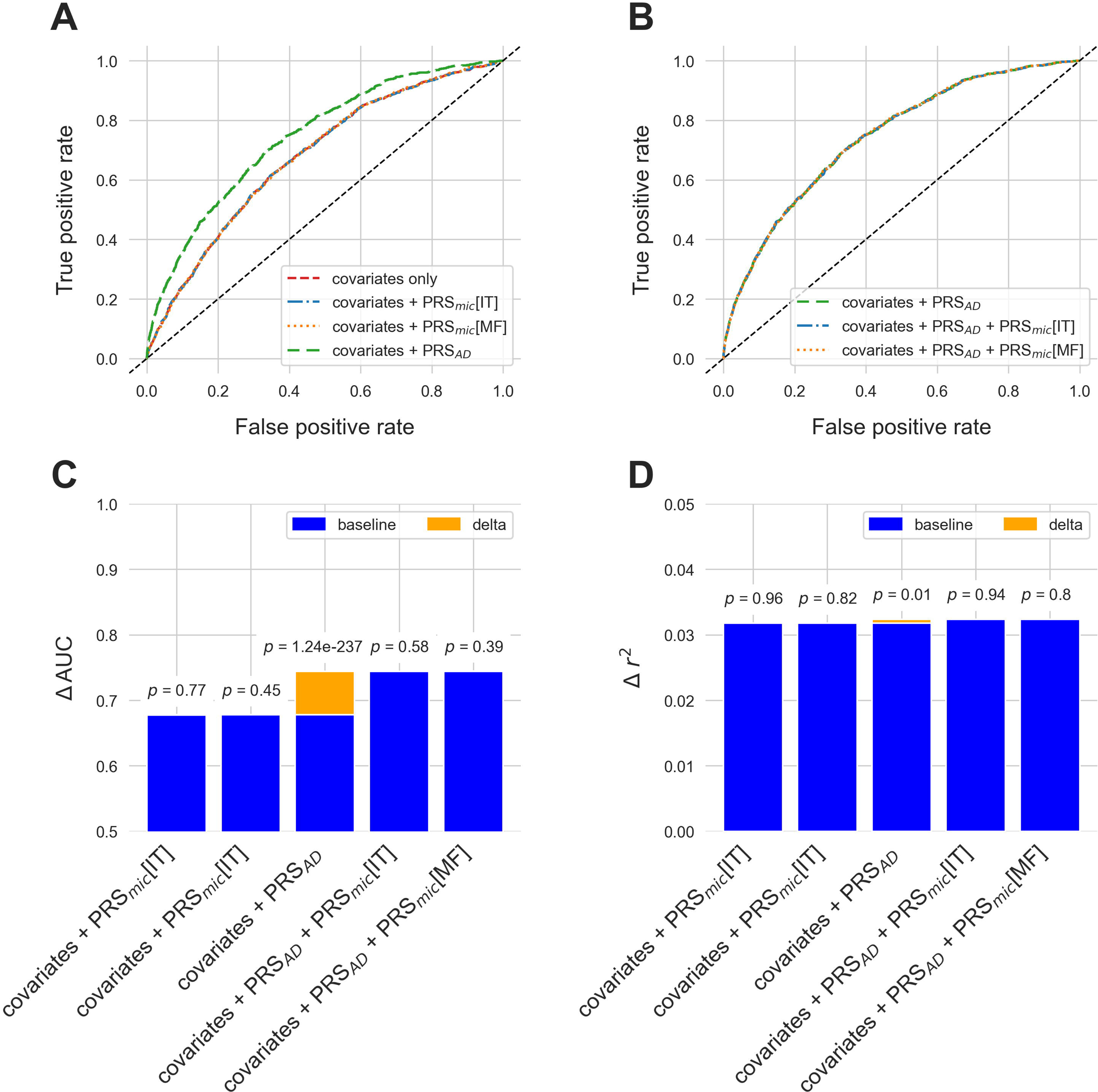
ROC curves for different models of Alzheimer’s disease diagnosis in the UK Biobank with **A)** the addition of PRS_mic_ and PRS_AD_ to covariates-only models, and **B)** the addition of PRS_mic_ to models including both covariates and PRS_AD_. Increase to AUC is presented in orange in **C)** with *p*-values from likelihood ratio tests indicated above each bar. Similarly, increase in variance explained for models of cognitive performance in CLSA are presented in **D)**. All scores used in these models were calibrated with age, sex, and education levels as covariates.

### Exploration of PRS associations with in vivo AD biomarkers in ADNI

Finally, we explored associations of both the PRS_AD_ and the PRS_mic_ with intermediate AD phenotypes in the ADNI sample, which had a wide array of available central and peripheral biomarker data available. First, our optimized PRS_AD_ was associated with worse cognitive performance on the ADAS-Cog 13-item scale (β=0.22, *p*=2.08×10^−17^). Higher levels of tau (β=0.26, *p*=2.42×10^−13^) and its phosphorylated variant (β=0.28, *p*=3.52×10^−15^) measured in CSF were also found to be associated with PRS_AD_. Lower levels of amyloid measured in both CSF (β=-0.30, *p*=8.57×10^−15^) and with PET tracer ^18^F-AV-45 in the inferior temporal region (β=-0.14, *p*=8.63×10^−4^) were associated with PRS_AD_ as well. Similarly, elevated levels of tau measured with PET tracer ^18^F-AV-1451 in both the inferior temporal (β=0.24, *p*=2.01×10^−5^) and midfrontal (β=0.18, *p*=1.05×10^−3^) regions were also associated with our score. We also observed associations with elevated neurofilament light chain (β=0.11, *p*=0.01) and decreased tumor necrosis factor alpha (β=-0.10, *p*=0.03) in plasma, as well as increased CSF neurofilament light chain (β=0.16, *p*=2.70×10^−3^). However, these did not survive correction for multiple testing. The only AD phenotype not observed to be associated with PRS_AD_ is levels of PET amyloid in the midfrontal region (*p*=0.17).

No association with any AD phenotype and PRS_mic_ survived correction for multiple testing. At uncorrected significance levels, the inferior temporal score, PRS_mic_[IT], is significantly associated with both CSF (β=0.09, *p*=0.02) and brain (inferior temporal PET: β=-0.11, *p*=7.44×10^−3^; midfrontal PET: β=0.08, *p*=0.05) amyloid. Significant associations were also found with cognition (β=-0.06, *p*=0.02) and plasma markers TNF-α (β=0.11, *p*=0.03) and NfL (β=0.11, *p*=0.02). PRS_mid_[MF] on the other hand, was nominally associated with all markers tested except for PET imaging markers for tau (inferior temporal PET: *p*=0.09; midfrontal PET, *p*=0.12). However, the direction of effect is inconsistent with the associations found with PRS_AD_ with the exception of plasma TNF-α (β=-0.13, *p*=5.12×10^−3^). Results for all the phenotypes tested, including those that did not pass correction for multiple testing are summarized in **Figure 2** and **Table S2**.

**Figure 2:**
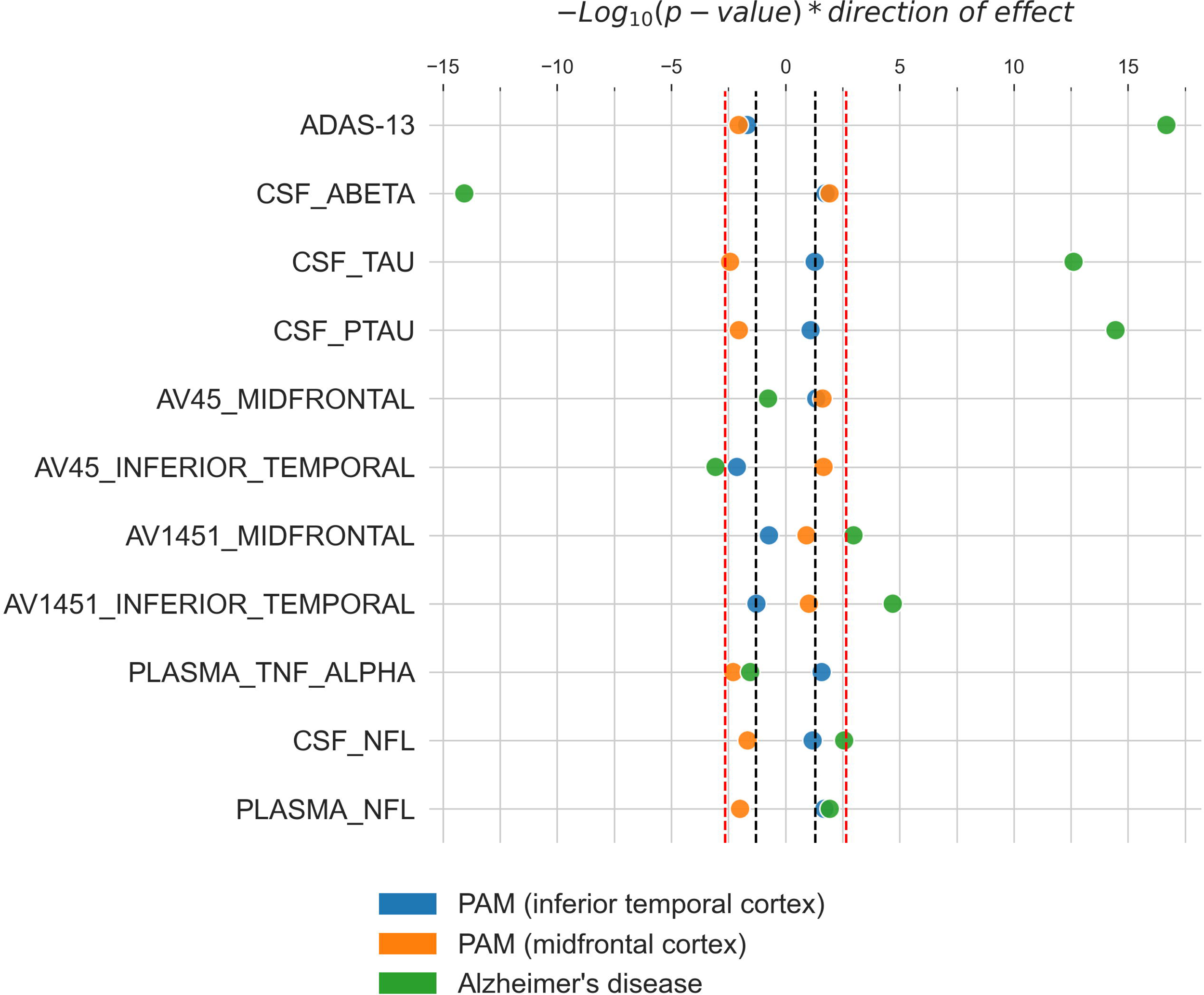
Associations of microglial-activation specific PRSs (IT, blue; MF, orange) and Alzheimer’s disease (AD) specific PRS (green) with AD-related phenotypes. −Log_10_(*p*-values), weighted by direction of effect, indicate the strength of evidence for association of each AD-related phenotype with the scores. The red dotted lines indicate corrected statistical significance thresholds, and the black dotted lines indicate uncorrected thresholds of *p*=0.05. All *p*-values are two-sided and calculated from linear regression models. Model covariates included age, sex, and education levels.

## Discussion

We developed and tested a novel PRS for morphological microglial activation as an adjunct to existing PRS as predictors of AD and cognitive performance in mid-late life. Such a score would contribute to our understanding of the genetics of neuroimmune mechanisms of pathological aging and potentially improve the clinical utility of genetic diagnostic and prognostic tools. We performed a rigorous resampling procedure using independent data and a proxy phenotype for microglial activation to identify optimal PRS hyperparameters, finding significant associations between these optimal scores and levels of TNF-α. We then investigated the utility of PRS_mic_ by testing for predictive ability in two external validation cohorts, the UK Biobank and CLSA. We observed no significant associations between the PRS_mic_ and either AD diagnosis or cognitive performance, nor any significant improvement in PRS_AD_ model performance with the inclusion of PRS_mic_.

Our largely negative results, especially in the inability of the PRS_mic_ to improve upon predictions from existing PRS for AD, point to four possible conclusions: 1) the genetic propensity for microglial activation is not related to the pathogenesis of AD, 2) the clumping and thresholding PRS methodology was unable to capture the genetic propensity for microglial activation, 3) our PRS_mic_ was not portable due to population and phenotypic measurement differences between the source GWAS, calibration, and optimization samples, and 4) the source GWAS for microglial activation was underpowered and did not produce durable estimates of SNP-based effects suitable for polygenic scoring. Here, we will address each of these possible conclusions.

First, microglial activation is a well-studied phenomenon that has been extensively characterized at multiple stages of AD pathogenesis and progression. The post-mortem morphological phenotype of PAM has also been well characterized at the clinical, cognitive, neuropathological, and molecular levels.^10^ What is less clear is whether or not genetic signals for this phenotype represent stable traits and mechanisms which hold predictive value for AD at the population level. In support of our endeavor, neuroinflammatory traits have been represented by genetic instruments previously with varying results.^40,41^

Second, the clumping and threshold method is only one of dozens of PRS methods available today. We chose this method due to 1) its optimal performance in the prediction of AD vs. other available methods,^37^ and 2) its simplicity and ease of interpretation. Also, unlike some other methods, clumping and thresholding requires the selection of hyperparameters that govern the handling of LD and the set of included SNPs, which can have dramatic impacts on PRS performance^37^. Calibration of these hyperparameters can be a major challenge, especially in the absence of independent datasets. Some very recent advances have been made in this space.^42^ In our case, the lack of direct measures of microglial activation outside of the source GWAS cohort created the need for a proxy phenotype in our calibration procedure. TNF-α was selected as it is a known peripheral marker of microglial activation,^43,44^ specifically as measured by morphology and measured physiological activity, which is most similar to the PAM phenotype. TNF-α is also known to be elevated in both the brains and plasma of AD patients, with post-mortem studies indicating proximity to amyloid plaques,^45^ further supported by studies of human microglial cell cultures showing induction of TNF-α release by amyloid plaques.^46^ Genome wide association studies have also consistently linked TNF-α receptor with AD. Associations with peripheral blood-based TNF-α, however, have yet to be fully elucidated in humans despite strong evidence from pre-clinical models.^47–49^ As more peripheral or *in vivo* imaging markers of PAM are identified, more accurate polygenic scores for microglial activation can be developed, eliminating the need of a proxy phenotype, such as TNF-α.

Third, portability is a known challenge with PRS development and is not entirely unexpected given the sensitivity of the scores to their method of construction.^50^ Our results serve to exemplify the challenges of indexing biological signals that are phenotypically relevant but difficult to ascertain, as is the case of neuroinflammation in Alzheimer’s disease. We have shown that genetic signals specific to microglial activation are associated with other measures of neuroinflammation. However, the effects observed in ADNI may be partially attributed to the similarity to ROS/MAP, the cohort where the proportion of activated microglia was originally characterized in. Our external validation cohorts, on the other hand, are community-based representative samples with no particular enrichment for late-life or Alzheimer’s disease, making establishing the link between neuroinflammation and AD more difficult.

Finally, the source GWAS that produced the summary statistics that our novel scores were based on only included up to 225 participants, which may not provide adequate statistical power.^51^ Nonetheless, our source GWASs identified two genome-wide significant hits, one of which was validated in an independent sample of TSPO PET imaging. And while microglial activation and proliferation are known to be sensitive to environmental exposures, the genetic basis for the PAM phenotype has been shown to genetically overlap with heritable risk for multiple traits and disorders, including Alzheimer’s disease and Crohn’s disease, further motivating investigations of its genetic causes. Another challenge with the source GWAS is that, due to the small sample size, formal estimates of SNP-based heritability were not reliable, meaning we cannot verify if the trait has a heritability above h2=0.05, which is an oft-used heuristic for determining the suitability of a trait for PRS modelling.^52^

Following best practices for rigorous polygenic risk score development, we did not find evidence to support the hypothesis that a PRS for morphological microglial activation can improve models of cognitive performance and AD in late life beyond a benchmark AD-specific PRS. While our study suggests that there may be some links between the genetic signals for morphological microglial activation and circulating TNF-a, we believe that larger, better-powered GWAS are required on such immune phenotypes prior to further testing.

## Data Availability

All data used in this study are publicly available upon registration and compliance with the data use agreements from each respective institute: the Alzheimer's Disease Neuroimaging Initiative (ADNI: https://adni.loni.usc.edu/), the UK Biobank (https://www.ukbiobank.ac.uk/), and the Canadian Longitudinal Study on Aging (CLSA: https://www.clsa-elcv.ca/).

https://www.ukbiobank.ac.uk/

https://www.clsa-elcv.ca/

## Acknowledgements

Data collection and sharing for this project was funded by the Alzheimer’s Disease Neuroimaging Initiative (ADNI) (National Institutes of Health Grant U01 AG024904) and DOD ADNI (Department of Defense award number W81XWH-12-2-0012). ADNI is funded by the National Institute on Aging, the National Institute of Biomedical Imaging and Bioengineering, and through generous contributions from multiple partners. The Canadian Institutes of Health Research is providing funds to support ADNI clinical sites in Canada. Private sector contributions are facilitated by the Foundation for the National Institutes of Health (www.fnih.org). The grantee organization is the Northern California Institute for Research and Education, and the study is coordinated by the Alzheimer’s Therapeutic Research Institute at the University of Southern California. ADNI data are disseminated by the Laboratory for Neuro Imaging at the University of Southern California. This research has been conducted using the UK Biobank Resource under Application Number 61530. This research was made possible using the data/biospecimens collected by the Canadian Longitudinal Study on Aging (CLSA) [Genome-wide Genetic Data Release version 3.0; Baseline Tracking Dataset version 4.0]. Funding for the Canadian Longitudinal Study on Aging (CLSA) is provided by the Government of Canada through the Canadian Institutes of Health Research (CIHR) under grant reference: LSA 94473 and the Canada Foundation for Innovation. ROSMAP is supported by P30AG10161, P30AG72975, R01AG15819, R01AG17917. U01AG46152, and U01AG61356.

## Conflict of interest

The author(s) declare(s) that there is no conflict of interest with respect to the research, authorship, and/or publication of this article.

## Funding

The author(s) disclosed receipt of the following financial support for the research, authorship, and/or publication of this article: the Koerner Family Foundation New Scientist Program, the Krembil Family Foundation, the Canadian Institutes of Health Research, and the Ontario Graduate Scholarship.

## Authors’ contributions

E.S.T.: conceptualization, methodology, software, formal analysis, writing - original draft, visualization. T.J.H.: resources, data curation, writing - review & editing. M.M.: data curation. D.A.B.: writing - review & editing. D.F.: conceptualization, methodology, writing - review & editing, supervision, funding acquisition.

## Ethics approval and consent to participate

The Alzheimer’s Disease Neuroimaging Initiative obtains participant consent at enrolment and is registered with an ICMJE-approved registry (ClinicalTrials.gov registry numbers: ADNI 1: <u>NCT00106899</u>; ADNI GO: <u>NCT01078636</u>; ADNI 2: <u>NCT0123197</u>; ADNI 3: <u>NCT02854033</u>). The UK Biobank has approval from the North West Multi-centre Research Ethics Committee (https://www.hra.nhs.uk/about-us/committees-and-services/res-and-recs/search-research-ethics-committees/north-west-haydock/) and has obtained informed consent from all participants. This study is approved by the UK Biobank under application ID 61530. The Canadian Longitudinal Study on Aging (CLSA) obtains consent at each data collection site and its associated research ethics board. The CLSA has approved a data access agreement for this study (Application Number: 2006026).

## Supplemental material

Supplemental material for this article is available online.

## Notes

### Competing Interest Statement

The authors have declared no competing interest.

### Author Declarations

The ethics committee of the Alzheimer's Disease Neuroimaging Initiative has given ethical approval for this work. The UK Biobank has approval from the North West Multi-centre Research Ethics Committee (https://www.hra.nhs.uk/about-us/committees-and-services/res-and-recs/search-research-ethics-committees/north-west-haydock/) and has obtained informed consent from all participants. This study is approved by the UK Biobank under application ID 61530. The Canadian Longitudinal Study on Aging (CLSA) obtains consent at each data collection site and its associated research ethics board. The CLSA has approved a data access agreement for this study (Application Number: 2006026).

