## Supplementary Data for "Testing a polygenic risk score for morphological microglial activation in Alzheimer’s disease and aging"

the Alzheimer’s Disease Neuroimaging Initiative**

1) Krembil Centre for Neuroinformatics, Centre for Addiction and Mental Health, Toronto, ON, CANADA

2) Institute of Medical Science, Temerty Faculty of Medicine, University of Toronto, Toronto, ON, CANADA

3) Vanderbilt Memory and Alzheimer's Centre, Vanderbilt University Medical Center, Nashville, TN, USA

4) Rush Alzheimer’s Disease Center, Rush University Medical Center, Chicago, IL., USA

5) Dalla Lana School of Public Health, University of Toronto, Toronto, ON, CANADA

6) Department of Psychiatry, Temerty Faculty of Medicine, University of Toronto, Toronto, ON, CANADA

*Corresponding author

Daniel Felsky PhD

Independent Scientist, Krembil Centre for Neuroinformatics, Centre for Addiction and Mental Health

Assistant Professor, Department of Psychiatry and Dalla Lana School of Public Health, University of Toronto

12th Floor, 250 College Street, Toronto ON, M5T 1R8, Canada

www.felskylab.com

(416) 535 8501 x33587

**Data used in preparation of this article were obtained from the Alzheimer’s Disease Neuroimaging Initiative (ADNI) database (adni.loni.usc.edu). As such, the investigators within the ADNI contributed to the design and implementation of ADNI and/or provided data but did not participate in analysis or writing of this report. A complete listing of ADNI investigators can be found at: <http://adni.loni.usc.edu/wp-content/uploads/how_to_apply/ADNI_Acknowledgement_List.pdf>

**Running Headline:** Microglia polygenic score for Alzheimer’s disease

**Table of Contents**

| **Table S1:** | Demographics of study participants included in analysis. | 3 |
| --- | --- | --- |
| **Table S2:** | Statistics from linear models of Alzheimer’s disease-related phenotypes with microglial-activation and Alzheimer’s disease specific PRSs as explanatory variables. | 5 |
| **Figure S1:** | Bootstrapped model performance as a function of 1) the clumping correlation threshold, 2) the clumping window size, and 3) the SNP inclusion threshold. Each point represents one linear model with a polygenic risk score term modeling plasma TNF-α (**A**, **B**, and **C**) and Alzheimer’s disease diagnosis (**D**, **E**, and **F**). All models control for age, sex, and education level. | 6 |
| **Figure S2:** | Bootstrapped model performance as a function of 1) the clumping correlation threshold, 2) the clumping window size, and 3) the SNP inclusion threshold. Each point represents one linear model with a polygenic risk score term modeling plasma TNF-α (**A**, **B**, and **C**) and Alzheimer’s disease diagnosis (**D**, **E**, and **F**). No covariates were included in the linear models. | 7 |
| **Figure S3:** | ROC curves for different models of Alzheimer’s disease diagnosis in the UK Biobank with **A)** the addition of PRS_mic_ and PRS_AD_ to covariates-only models, and **B)** the addition of PRS_mic_ to models including both covariates and PRS_AD_. Increase to AUC is presented in orange in **C)** with *p*-values from likelihood ratio tests indicated above each bar. Similarly, increase in variance explained for models of cognitive performance in CLSA are presented in **D)**. All scores used in these models were calibrated without covariates. | 8 |

**Table S1:** Demographics of study participants included in analysis.

| **Phenotype** | | **Missing** | **Total** | | **Alzheimer’s disease** | | **Cognitively normal** | | **Statistic* (*p*-value)** |
| --- | --- | --- | --- | --- | --- | --- | --- | --- | --- |
|  |  |  | **Count** | **Mean (SD)** | **Count** | **Mean (SD)** | **Count** | **Mean (SD)** |  |
| **Alzheimer’s Disease Neuroimaging Initiative (ADNI)** | | | | | | | | | |
| Alzheimer’s disease diagnosis | | 441 | 973 | - | 494 | - | 479 | - | - |
| Sex | | 0 | 1414 | - | - | - | - | - | 15.86 (6.81x10^-5^)* |
|  | Male | 263 | 523 | - | 297 | - | 226 | - | - |
|  | Female | 178 | 450 | - | 197 | - | 253 | - | - |
| Age (years) | | 0 | 1414 | 73.67 (7.12) | 494 | 74.66 (7.36) | 479 | 72.60 (6.36) | 4.66 (3.54x10^-6^) |
| Education (years) | | 0 | 1414 | 16.07 (2.78) | 494 | 15.51 (2.94) | 479 | 16.70 (2.43) | -6.85 (1.33x10^-11^) |
| ADAS-Cog | | 10 | 1404 | 15.36 (8.94) | 487 | 23.34 (8.60) | 476 | 8.66 (4.18) | 33.58 (3.72x10^-164^) |
| CSF β-amyloid (pg/ml) | | 777 | 637 | 838.41 (366.07) | 290 | 672.10 (279.06) | 128 | 1055.25 (373.12) | -11.62 (3.26x10^-27^) |
| CSF tau (pg/ml) | | 654 | 760 | 286.09 (118.46) | 303 | 343.44 (122.58) | 183 | 232.24 (80.39) | 10.93 (5.21x10^-25^) |
| CSF phosphorylated tau (pg/ml) | | 654 | 760 | 27.54 (13.3) | 303 | 34.34 (14.07) | 183 | 21.09 (8.28) | 11.59 (1.46x10^-27^) |
| AV45 midfrontal (SUVR) | | 837 | 577 | 4.67 (0.33) | 131 | 4.62 (0.29) | 214 | 4.65 (0.34) | -0.63 (0.53) |
| AV45 inferior temporal (SUVR) | | 837 | 577 | 2.26 (0.19) | 131 | 2.22 (0.16) | 214 | 2.28 (0.21) | -3.13 (0.002) |
| AV1451 midfrontal (SUVR) | | 1138 | 276 | 4.36 (0.91) | 25 | 5.66 (2.43) | 195 | 4.20 (0.30) | 8.04 (5.83x10^-14^) |
| AV1451 inferior temporal (SUVR) | | 1138 | 276 | 2.54 (0.51) | 25 | 3.49 (1.04) | 195 | 2.41 (0.19) | 13.07 (3.24x10^-29^) |
| CSF NfL (ng/L) | | 1088 | 326 | 1474.13 (1060.54) | 185 | 1590.83 (1171.12) | 60 | 1087.57 (405.46) | 3.26 (0.001) |
| Plasma NfL (pg/ml) | | 951 | 463 | 42.3 (24.86) | 255 | 45.56 (22.59) | 111 | 33.78 (18.34) | 4.84 (1.90x10^-6^) |
| Mean plasma TNF-α (pg/mL) | | 964 | 450 | 0.85 (0.21) | 267 | 0.85 (0.22) | 42 | 0.86 (0.25) | -0.27 (0.78) |
| **UK Biobank** | | | | | | | | | |
| Alzheimer’s disease diagnosis | | 0 | 200924 | - | 2728 | - | 198196 | - | - |
| Sex | | 0 | 200924 | - | - | - | - | - | 0.74 (0.39)* |
|  | Male | 0 | 95099 | - | 1314 | - | 93785 | - | - |
|  | Female | 0 | 105825 | - | 1414 | - | 104411 | - | - |
| Age (years) | | 0 | 200924 | 64.13 (2.85) | 2728 | 65.73 (2.70) | 198196 | 64.11 (2.85) | 29.53 (3.42x10^-191^) |
| Education (years) | | 2624 | 198300 | 13.71 (5.34) | 2660 | 12.49 (5.39) | 195640 | 13.72 (5.34) | -11.78 (4.81x10^-32^) |
| **Canadian Longitudinal Study on Aging (CLSA)** | | | | | | | | | |
| Sex | | 0 | 11313 | - | - | - | - | - | - |
|  | Male | 0 | 5686 | - | - | - | - | - | - |
|  | Female | 0 | 5627 | - | - | - | - | - | - |
| Cognitive performance | | 0 | 11313 | 99.94 (15.15) | - | - | - | - | - |
| Age (years) | | 0 | 11313 | 69.63 (6.94) | - | - | - | - | - |
| Education (years) | | 0 | 11313 | 7.19 (2.45) | - | - | - | - | - |

ADAS-Cog = The Alzheimer's Disease Assessment Scale-Cognitive Subscale, CSF = cerebrospinal fluid, SUVR = standardized uptake value ratio, NfL = neurofilament light chain, TNF-α = tumour necrosis factor alpha

*The chi-square test for independence, corrected for the Yates’ continuity, was used to compare distributions of count data (i.e. sex), and the two-sided t-test was used for continuous data. The chi-squared statistic is reported for the former, and t statistic for the latter.

**Table S2:** Statistics from linear models of Alzheimer’s disease-related phenotypes with microglial-activation and Alzheimer’s disease specific PRSs as explanatory variables.

| Alzheimer’s disease-related phenotypes | PRS_AD_ | | PRS_mic_[IT] | | PRS_mic_[MF] | |
| --- | --- | --- | --- | --- | --- | --- |
|  | Beta | *p*-value | Beta | *p*-value | Beta | *p*-value |
| ADAS-Cog | 0.22 | **2.08x10^-17^** | -0.06 | 0.02 | -0.07 | 8.84x10^-3^ |
| β-amyloid (CSF) | -0.30 | **8.57x10^-15^** | 0.09 | 0.02 | 0.10 | 0.01 |
| Tau (CSF) | 0.26 | **2.42x10^-13^** | 0.07 | 0.05 | -0.10 | 3.74x10^-3^ |
| Phosphorylated tau (CSF) | 0.28 | **3.52x10^-15^** | 0.06 | 0.08 | -0.10 | 9.06x10^-3^ |
| AV45 (Midfrontal) | -0.06 | 0.17 | 0.08 | 0.05 | 0.10 | 0.02 |
| AV45 (Inferior Temporal) | -0.14 | **8.63x10^-4^** | -0.11 | 7.44x10^-3^ | 0.10 | 0.02 |
| AV1451 (Midfrontal) | 0.18 | **1.05x10^-3^** | -0.08 | 0.19 | 0.09 | 0.12 |
| AV1451 (Inferior Temporal) | 0.24 | **2.01x10^-5^** | -0.12 | 0.05 | 0.10 | 0.09 |
| NfL (CSF) | 0.16 | **2.70x10^-3^** | 0.10 | 0.06 | -0.13 | 0.02 |
| NfL (plasma) | 0.11 | **0.01** | 0.11 | 0.02 | -0.12 | 0.01 |
| Mean TNF-α (plasma) | -0.10 | **0.03** | 0.11 | 0.03 | -0.13 | 5.12x10^-3^ |

AD = Alzheimer’s disease, IT = inferior temporal, MF = midfrontal, ADAS-Cog = The Alzheimer's Disease Assessment Scale-Cognitive Subscale, CSF = cerebrospinal fluid, NfL = neurofilament light chain, TNF-α = tumour necrosis factor alpha

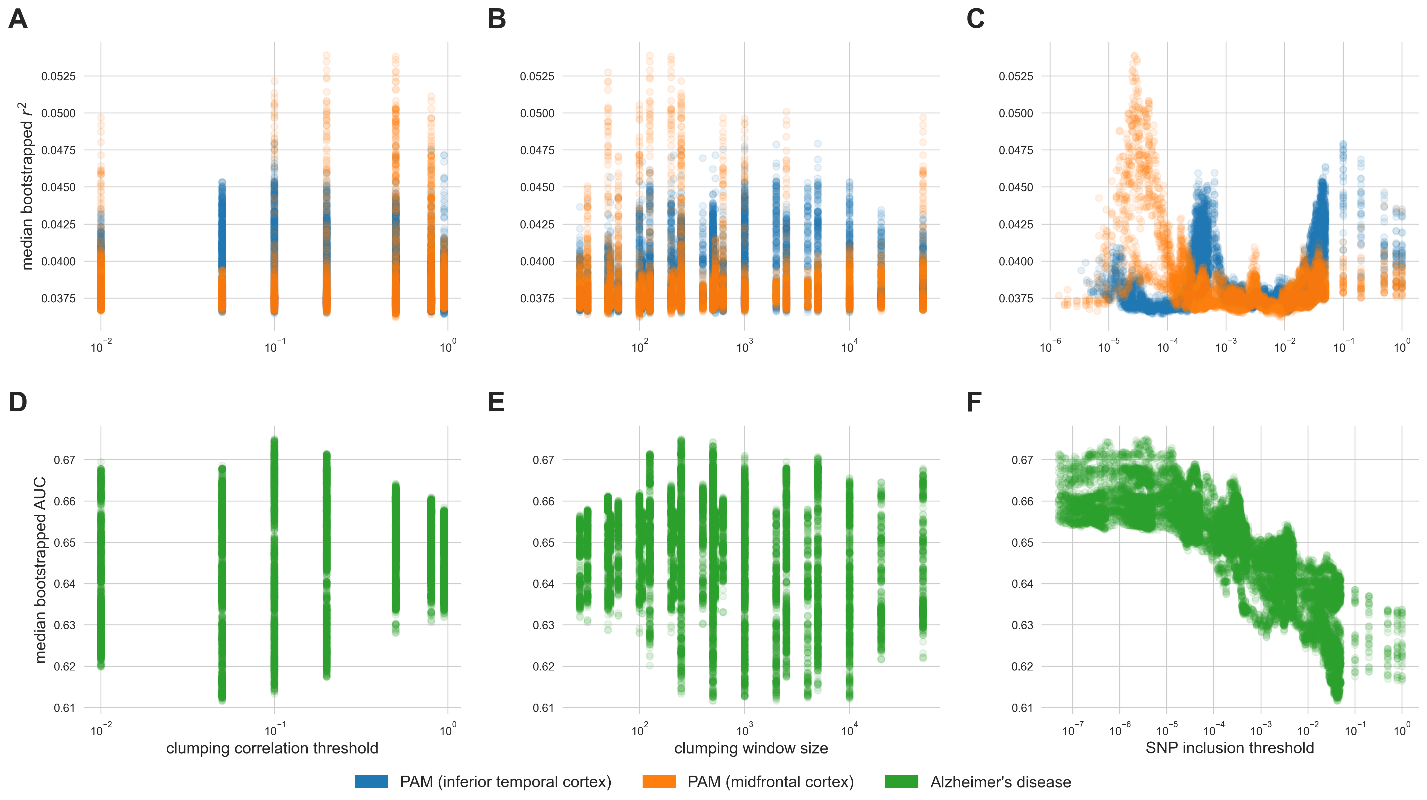

**Figure S1:** Bootstrapped model performance as a function of 1) the clumping correlation threshold, 2) the clumping window size, and 3) the SNP inclusion threshold. Each point represents one linear model with a polygenic risk score term modeling plasma TNF-α (**A**, **B**, and **C**) and Alzheimer’s disease diagnosis (**D**, **E**, and **F**). All models control for age, sex, and education level.

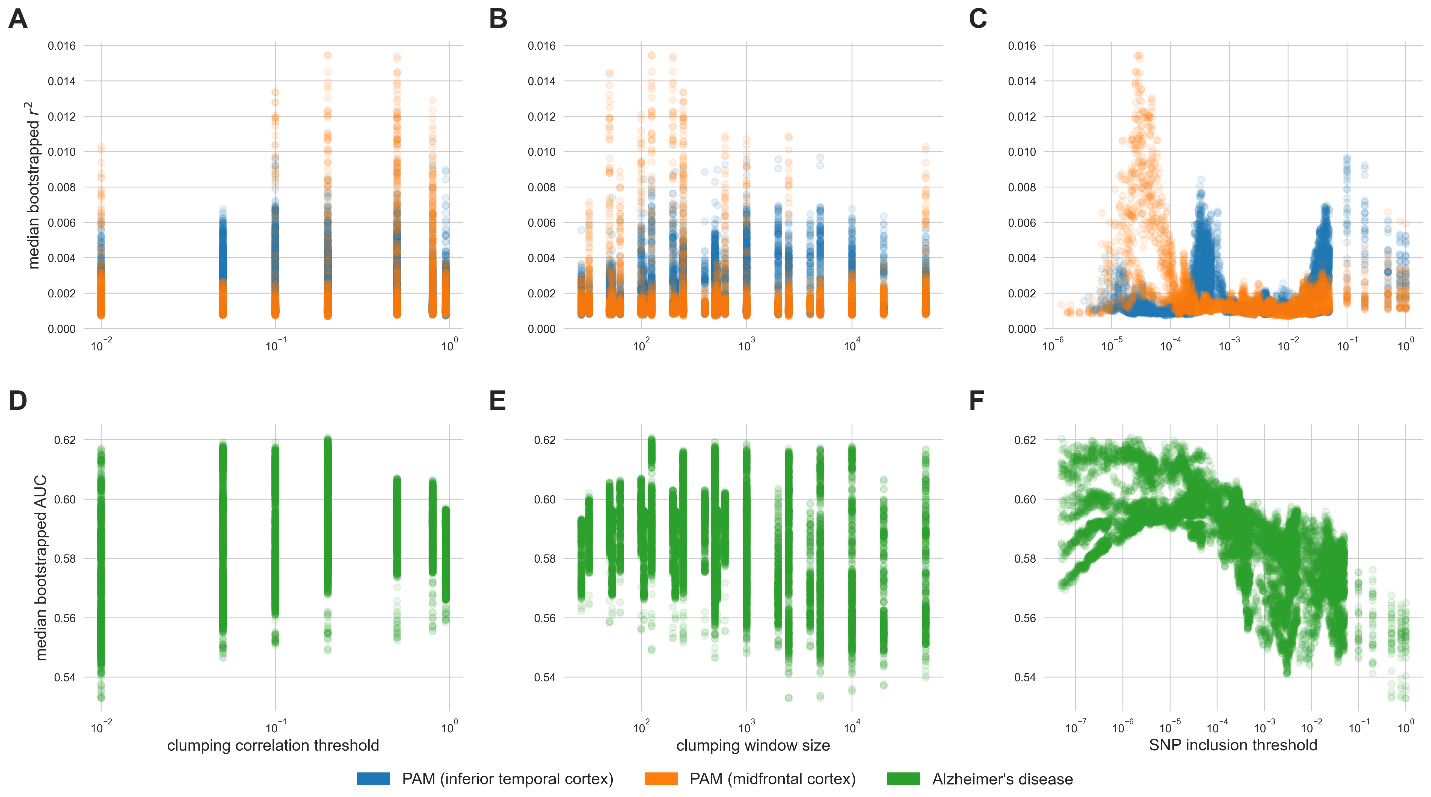

**Figure S2:** Bootstrapped model performance as a function of 1) the clumping correlation threshold, 2) the clumping window size, and 3) the SNP inclusion threshold. Each point represents one linear model with a polygenic risk score term modeling plasma TNF-α (**A**, **B**, and **C**) and Alzheimer’s disease diagnosis (**D**, **E**, and **F**). No covariates were included in the linear models.

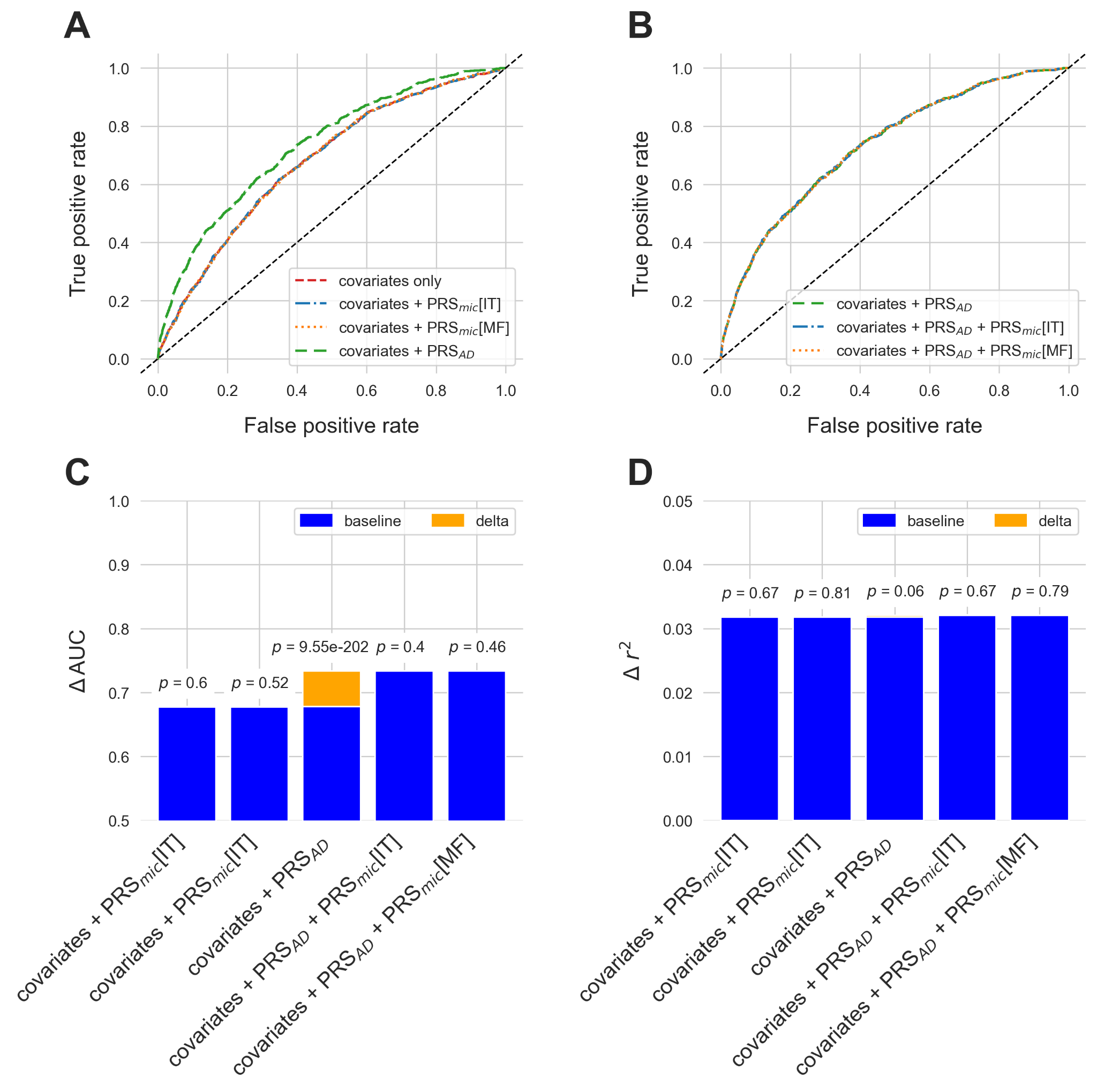

**Figure S3:** ROC curves for different models of Alzheimer’s disease diagnosis in the UK Biobank with **A)** the addition of PRS_mic_ and PRS_AD_ to covariates-only models, and **B)** the addition of PRS_mic_ to models including both covariates and PRS_AD_. Increase to AUC is presented in orange in **C)** with *p*-values from likelihood ratio tests indicated above each bar. Similarly, increase in variance explained for models of cognitive performance in CLSA are presented in **D)**. All scores used in these models were calibrated without covariates.
